# Biochemical and Immunological Predictors of Non-healing in Individuals with Early-stage Diabetic Foot Ulcers

**DOI:** 10.1101/2021.11.09.21266108

**Authors:** Jayashree Vijaya Raghavan, Shruthi Ksheera Sagar, Vinod Kumar Dorai, Rebecca Diya Samuel, Priyanka Arunachalam, H C Chaluvanarayana, Pavan Belahalli, S R Kalpana, Siddharth Jhunjhunwala

## Abstract

**Objective:** The goal of this study was to identify biochemical and immunological parameters from the blood as predictors of non-healing in early-stage diabetic foot ulcers.

**Research Design and Methods:** We performed a cross-sectional prospective cohort study among individuals with early-stage foot ulcers visiting the Karnataka Institute of Endocrinology Research over a 2.5-year period. Histopathological, biochemical, and immunological data (a total of 31 parameters) from 52 individuals were collected and analyzed to determine if predictors of non-healing may be identified. Data analysis was performed using traditional univariate analyses as well as univariate and multivariable logistic regression.

**Results:** Individual histopathological and biochemical parameters did not show any differences between healed and non-healed individuals. However, conventional univariate analysis and univariate logistic regression analysis showed that the expression of the cell-surface proteins CD63, HLA-DR and CD11b on monocytes (CD14^+^) was significantly lower in non-healed individuals, but with moderate discriminative ability as assessed by area under the curve (AUC) of Receiver Operating Characteristics (ROC) curve. In comparison, a multivariable logistic regression model identified four of the 31 parameters to be salient predictors and demonstrated high discrimination ability with an AUC of ROC value of 0.87. Among the four identified parameters, LDL cholesterol (OR 18.83, CI 18.83-342) and cell-surface expression of CD63 on monocytes (OR 0.12, CI 0.12-0.45) were significant.

**Conclusion:** Through this study we conclude that LDL cholesterol and cell-surface expression of CD63 on monocytes are strong positive and negative predictors of non-healing, respectively, in individuals with early-stage DFU. Following validation in a larger cohort, these parameters may be used by the clinician for early identification of non-healers.

## INTRODUCTION

Diabetic foot ulcers (DFU) are chronic wounds that exhibit delayed healing in diabetic individuals. About 19-34% of diabetic individuals are reported to be at risk of development of DFU(1). Currently, these wounds are managed through debridement, dressing, pressure off-loading, glycemic control and patient education(2). Treatment strategies include topical application of antibiotics and antimycotics to prevent infection, growth factor ointments to enhance cell proliferation for wound healing and negative pressure therapy for wound fluid drainage(2–4). Although a large proportion of early-stage wounds heal with timely care, a small but significant proportion of wounds fail to heal resulting in progression to gangrene formation necessitating limb amputation(3,5,6). Hence, an early-stage predictor for wounds that are unlikely to heal would help the clinician alter treatment strategies, which might reduce the chance of amputation.

Measurable DFU characteristics such as wound size and depth have been described as potential predictors of non-healing in later stage and amputated wounds(7–9). In addition, it has been suggested that decreased wound area at early times (such as 1 and 4 weeks) correlated with poor healing at later time points (12 weeks)(8). However, precise cutoffs for wound size or depth, and wound area reduction are difficult to establish as it may vary between study cohorts, ulcer stages and ulcer types(8,9). Chronic local inflammation (at the wound site) has also been correlated to delayed healing(3,10). Some of the parameters associated with inflammation in DFUs include excessive myeloid cell infiltration(11), increased levels of pro-inflammatory mediators(12), presence of neutrophil extracellular traps (NETs)(13,14), and increased levels of MMP9 (15). However, quantification of such ulcer parameters is often difficult in many clinical centers. A few studies also suggest presence of chronic low-grade systemic inflammation in addition to local inflammation in DFU individuals(16,17), quantification of which are possible in most clinics. One such cohort study by Dinh and colleagues showed that non-healed individuals had increased (as compared to individuals whose ulcers healed) serum levels of tumor necrosis factor-α (TNF-α), monocyte chemoattractant protein-1 (MCP-1), matrix metallopeptidase-9 (MMP-9), and fibroblast growth factor-2 (FGF-2) about 8 months prior to the development of ulcers(18). We conjectured that as myeloid cells play a vital role in the secretion of these cytokines and the establishment of inflammation, they might contribute to slower or absence of healing. However, their role in the healing of early-stage ulcers remains poorly characterized. The goal of the current study was to characterize the phenotype of myeloid cells in individuals with active early-stage ulcers and determine if their phenotype along with other clinical parameters may be used to predict non-healing of ulcers.

## METHODS

### Ethics statement

The study received approval from the institutional review board at the Karnataka Institute of Endocrinology Research (protocol number: IEC-KIER/04/28.10.2017). All procedures were conducted in accordance with the approved protocol.

### Recruitment, Sample Collection, and Follow-Up

Blood and wound biopsy samples were collected from individuals after obtaining informed consent at Karnataka Institute of Endocrinology and Research (KIER), Bengaluru, Karnataka, India. Stage of ulcer was graded by the clinician following Wagner’s grade of ulcer classification(19). Diabetic neuropathic patients who presented with a non-ischemic and uninfected early-stage (stage I and II) foot ulcer were selected for the study (age range 40 – 70 years). Patients presenting any associated complications such as significant cardiac or renal ailments were excluded from the study. A total of 83 patients meeting patient inclusion criteria provided informed consent and were recruited into the study. Among these, 31 individuals dropped out from the study primarily because they did not show up for follow-up visits. Data from 52 individuals, for whom follow-up was performed twice a week for at least one month, were used for further analysis.

Wound biopsy sample was collected before the debridement procedure, on the day of recruitment right before initiation of treatment. Biopsy samples were stored in 10% buffered formalin until further processing. Peripheral venous blood (10 ml) was collected from enrolled patients within 7 days of biopsy collection and used for biochemical testing and immunophenotyping. Patient’s wounds were dressed regularly. Wound healing was monitored for a period of one month by capturing images using a near infra-red based camera (WoundZoom, USA) at the time of dressing.

### Image analysis

Wound images captured by a near infra-red based camera were analyzed using ImageJ. Percentage reduction in wound area was calculated as a measure of healing. Threshold of 50% reduction in wound area on Day 30 was used to classify patients as healed.

### Histopathological analysis

Grossing was performed on biopsy samples stored in formalin prior to processing to identify site of ulcer and size. Dissected ulcerated tissue was then embedded in paraffin and sectioned. H&E stained slides were evaluated by a pathologist, who was blinded to the stage and healing status of the patients. The pathologist assessed epidermal and vascular anomalies, inflammatory cell infiltration, granulation tissue, collagen, and fibrin deposition.

### Biochemical parameter measurements

Biochemical measurements were performed immediately following blood collection at the clinical laboratory in KIER. Tests included blood glucose profile, complete hemogram, total lipid profile, urine profile, thyroid profile, total liver function test, and few other additional parameters such as serum creatinine, serum electrolytes and C-reactive protein. Salient biochemical parameters of interest were chosen for further analysis.

### Immunophenotyping

Peripheral venous blood (5 ml) was transported to the laboratories at Indian Institute of Science (IISc), where it was processed for further analysis (within 3 hours of blood collection to ensure analysis of granulocyte populations). Blood was centrifuged at 500 RCF for 5 minutes to separate plasma from cells. Plasma was stored in −80 °C, while the pellet was subjected to RBC lysis using Ammonium-Chloride-Potassium (ACK) lysis buffer at 1:12.5 (blood to lysis buffer) ratio by volume. Lysis was performed at room temperature (RT) for 10 minutes and quenched using three times the volume using 1X PBS buffer containing 4mM EDTA. Samples were centrifuged at 400 RCF for 4 minutes at 4 °C. Supernatant was decanted and pellet was re-suspended in 1 mL of 1X PBS EDTA buffer. An aliquot of the sample was stained with BD Horizon™ Fixable Viability Stain 510 to stain dead cells at a concentration of 0.3 μl/100 μl volume of 1 million cell suspension. Samples were incubated in dark at RT for 20 minutes before quenching with 1 mL of 1X PBS containing 1% BSA and 4mM EDTA (staining buffer). Cells were then fixed using 2% paraformaldehyde for 30 minutes at 4 °C. Samples were washed and re-suspended in 1mL staining buffer for staining with fluorophore conjugated antibodies procured from BD Biosciences.

Staining was performed according to manufacturer’s recommended dilutions. Antibodies used are summarized in Supplementary Table 1. Samples were quenched with 1 mL staining buffer following staining, washed, and re-suspended in 300 μl buffer for acquisition on flow cytometer (BD FACSCelesta™ Cell Analyzer, BD Biosciences, USA). Appropriate single-color controls were prepared using compensation beads (BD Biosciences, USA), which were stained with antibodies following manufacturer’s protocol. Additionally, fluorescence minus one (FMO) controls were prepared to correct for group effect of fluorophores on spectral spill. Briefly, live-dead dye stained cells were stained with antibodies for all colors except one which was replaced by respective isotype to prepare FMO for a color of interest. Similar controls were prepared for all fluorophores mentioned in Supplementary Table 1 and used for intensity correction at the time of data analysis. Voltages were set on system using compensation beads at the time of sample acquisition and a minimum of 100,000 CD45^+^ live events were acquired using BD FACSDiva™ Version 6 Software system (BD Biosciences, USA).

### Flow cytometry data analyses

All data analyses were performed on FCS files using FlowJo™ v10.6 (Becton Dickinson, USA). Compensation was performed on FlowJo using compensation beads. CD14 and CD15 were used to identify monocytes and granulocytes, respectively. FMO’s were used to draw appropriate gates. A threshold of minimum 100 events was set to report percentage positive cells and median fluorescence intensity (MFI) values for all markers expressed by CD14^+^ and CD15^+^ cells. Corresponding FMO values of markers were subtracted to correct for group effect of fluorophores on spectral spill. Corrected MFI values were tabulated for further analyses.

### Data preprocessing and logistic regression model

Biochemical and immunological data of all patients was combined into one matrix and standardized prior analysis. Standardization was performed by centering data around mean and dividing each observation by standard deviation. Univariate and multivariable logistic regression models were built using standardized data using *glm* function in *stats* package of R. Stepwise feature selection was performed using packages *MASS*(20) and *magrittr*(21) with low Akaike’s information criterion (AIC) score as selection criterion for identifying salient predictors. Unadjusted odd’s ratio was reported with 95% confidence interval for multivariable logistic regression model. ROC analysis was performed using *pROC* package in R(22). ROC plots were created using the same package for univariate and multivariable logistic regression models.

### Statistics

Fisher’s exact test was performed to analyze categorical histology data in R using *stats* package. Heatmap was created using *ComplexHeatmap* package in R(23). Univariate analysis of all predictors was performed using student’s t-test with Welch’s correction using GraphPad Prism 8. Wald’s test for significance of predictors was performed by *glmnet* package as part of the routine. Model evaluation statistics such as Chi-square goodness of fit (GOF) test and Hosmer-Lemeshow (HL) test was performed using *stats* and *ResourceSelection* (24) package in R respectively. AIC score was obtained for all models using *stats* package. Model fit plots were created using *ggplot2* package in R(25). RStudio 4.1.0 was used for all R based analyses. Raw data and R scripts used for performing analyses are uploaded on https://github.com/Immunoengineeringlab/DFU_IISc_KIER_JVR_Data.

## RESULTS

### Cohort characteristics

The final study cohort comprised of 52 diabetic individuals, with either Stage I (30) or Stage II (22) foot ulcers. Their clinical characteristics are summarized in Table 1. We observe an underrepresentation of females in the study (only 8 females in comparison to 44 males), which could be due to sociological reasons. Through regular follow-up for one month that involved standard clinical care and imaging of the wound (Supplementary Figure 1), we determined that the ulcers had healed (>50% reduction in wound area) in 33 individuals, while it remained non-healed in 19 individuals. Our healing data shows that ulcers healed in 66.67% of stage I and 59% of stage II foot ulcer individuals with the standard care provided at our clinic.

**Table 1:** Clinical and blood sugar characteristics of recruited individuals. Data from 30 individuals with stage I and 22 individuals with stage 2 ulcers is presented here. LCI and UCI indicate lower and upper confidence intervals, respectively.

|  | Stage of ulcer | Median | LCI | UCI |
| --- | --- | --- | --- | --- |
| **Age** | I | 52.50 | 49.47 | 55.53 |
|  | II | 54.50 | 50.55 | 58.45 |
| **BMI** | I | 25.54 | 24.15 | 26.93 |
|  | II | 27.26 | 25.67 | 28.84 |
| **HbA1c %** | I | 9.35 | 8.72 | 9.98 |
|  | II | 9.30 | 8.71 | 9.89 |
| **FBS (mg/dl)** | I | 155.50 | 140.02 | 170.98 |
|  | II | 147.00 | 123.87 | 170.13 |
| **PPBS (mg/dl)** | I | 217.00 | 192.10 | 241.90 |
|  | II | 221.00 | 186.06 | 255.94 |

### Histopathological analysis of wound biopsy

Hematoxylin and Eosin (HE) stained sections of the ulcer biopsies were analyzed by a pathologist to assess the damage to skin tissue and levels of inflammatory cell infiltration. The pathologists report revealed that while presence of granulation tissue correlated with stage of ulcer, there was an absence of correlation between any of the histological features and healing of ulcers (Supplementary Figure 2A and 2B). This suggests that histopathological parameters may not be good predictors of ulcer healing.

### Univariate analysis of biochemical and immunological parameters

We also collected peripheral venous blood from the recruited individuals for biochemical analysis and immunological characterization of neutrophils and monocytes in circulation. Immunological characterization was performed using flow cytometry (Supplementary Figure 3A), and expression levels of various cell surface proteins are reported as the median fluorescence intensity (MFI) (Supplementary Figure 3B).

The standardized numerical values of the data collected from both biochemical analysis and immunological characterization (parameters) are presented as a heat-map in Figure 1. A clear separation among parameters was not observed between healed and non-healed individuals. Nevertheless, a univariate analysis was performed using Student’s t-test with Welch’s correction on each parameter, and this analysis revealed that none of the biochemical parameters showed significant differences among the healed and non-healed individuals (Supplementary Table 2). However, the analysis did show that the expression levels of three proteins on monocytes (CD14 expressing cells), CD63, HLA-DR and CD11b, were significantly higher on individuals whose ulcers had healed as compared to those whose ulcers had not healed (Supplementary Table 2 and Supplementary Figure 4). When a receiver operator curve (ROC) analysis was performed (Supplementary Table 3) to assess the discriminative capability of each parameter, all three were observed to have relatively moderate area under the curve (AUC) values and were deemed to be not effective in discriminating between non-healed ulcers and healed ulcers. Hence, we next used logistic regression, a binary classifier model to assess outcome prediction.

**Figure 1:**
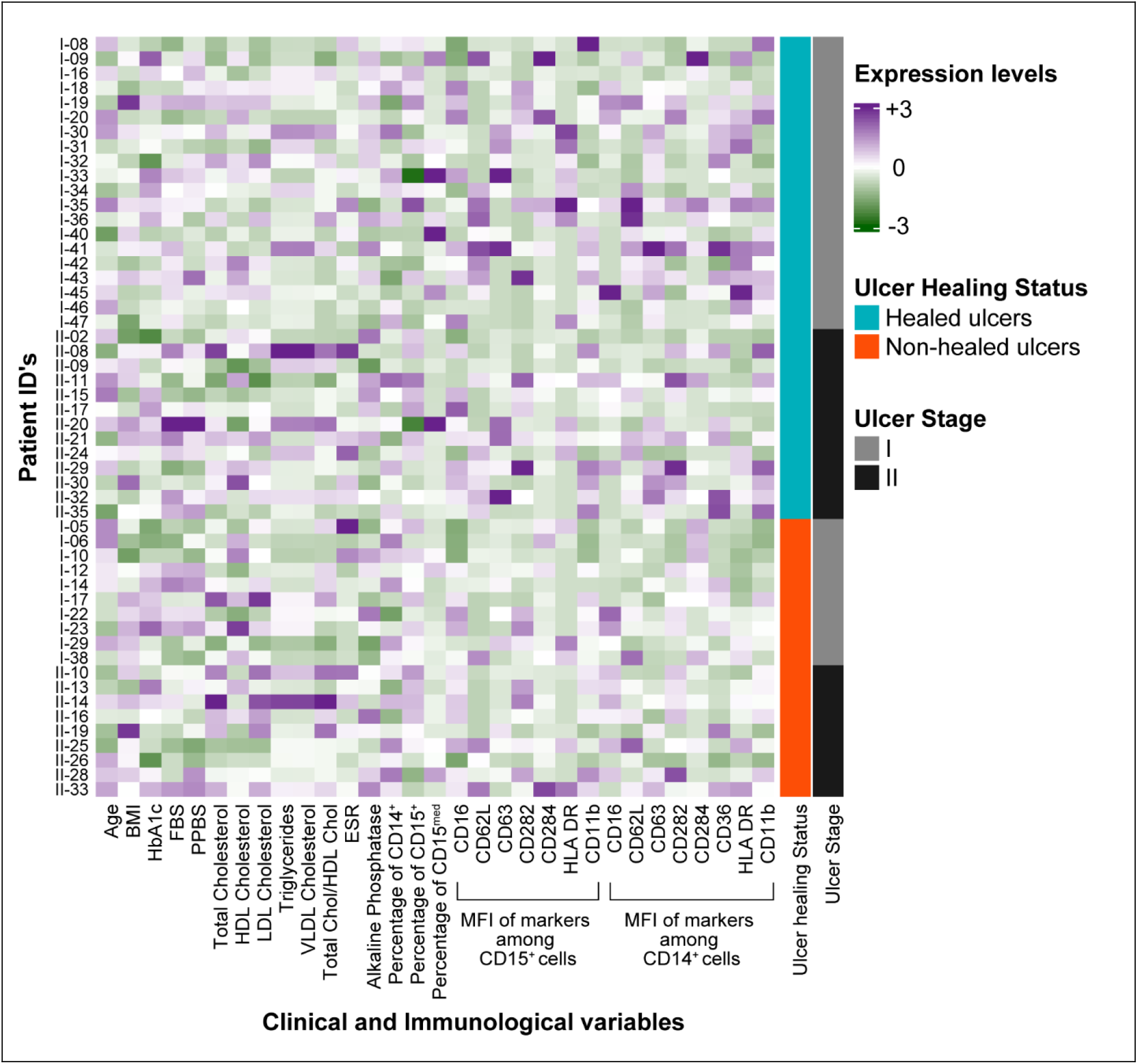
Heatmap of standardized biochemical and immunological parameters measured in 52 individuals with diabetic foot ulcer. Standardized data of each parameter is represented as heatmap with intensities varying between brown (−3), white (0) and purple (+3). Annotation – *Ulcer healing status* indicates if the individual was classified as healed or non-healed after 1 month follow-up, and *Ulcer stage* represents the stage of ulcer in each individual.

### Univariate logistic regression analysis

A univariate logistic regression analysis was performed to evaluate predictive capability of the three individual parameters identified above. In this analysis, the parameters MFI of CD63 and HLA-DR among monocytes (CD14^+^) were observed to be significant according to the Chi-square Goodness of Fit (GOF) test, and only MFI of CD63 among monocytes (CD14^+^) was significant when evaluated using the Wald’s test (Supplementary Table 4). Additionally, the odd’s ratio (OR) for MFI of CD63 among monocytes (CD14^+^) was found to be 0.26 (CI 0.08-0.68) suggesting strong negative association with healing outcome. The univariate logistic regression model’s prediction against actual outcomes for each univariate model are shown in Figure 2A. Further, ROC analysis was performed to evaluate the discriminative capability of univariate logistic regression models (Figure 2B). AUC was found to be 0.73 for CD63, 0.65 for HLA-DR and 0.63 for CD11b expressing monocytes indicating that the discriminative ability of the univariate logistic regression models was not better than the traditional univariate analysis. Hence, we next performed a multivariable logistic regression analysis.

**Figure 2:**
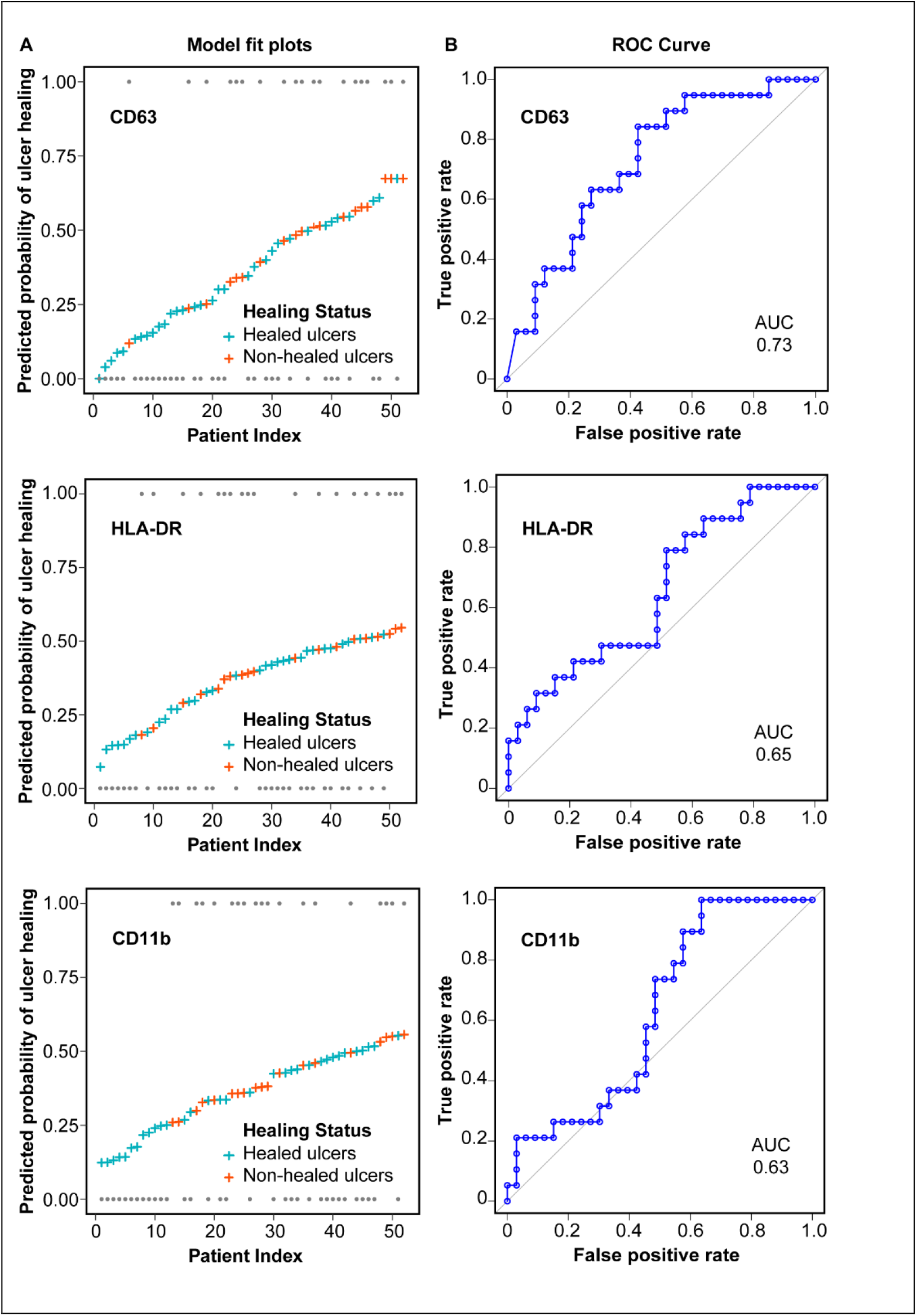
Univariate logistic regression model. **A** – The predicted probabilities CD63, HLA-DR and CD11b are shown (blue – healed; orange – non-healed) against reference probabilities indicated by grey dots. **B** – Receiver operator characteristics (ROC) curve of CD63, HLA-DR and CD11b has been plotted. Corresponding area under the curve was observed to be 0.73, 0.65 and 0.63, respectively.

### Multivariable logistic regression analysis

To determine if a combination of parameters enhances model performance, two multivariable logistic regression models were built initially. One model included only the biochemical and cellular parameters while the other included only immunological phenotypic parameters. Stepwise feature selection was performed to identify and retain only salient predictors to build the model. For the model including biochemical and cellular parameters, among the 16 measured parameters six salient parameters were identified by stepwise feature selection technique, four of which were significant according to Wald’s test (Table 2). The model performance was found to be significant based on a Chi-square goodness of fit (GOF) test (p = 0.011). A Hosmer-Lemeshow (HL) goodness of fit test was also performed to evaluate agreement between model’s predicted and expected event rates across deciles of risk groups. HL test p value was found to be 0.411 implying lack of evidence for disagreement between predicted and expected event rates. Additionally, Akaike Information Criterion (AIC), an estimator of out-of-sample prediction error to assess model performance was found to be 65.74. Odds ratio for each predictor along with CI is summarized in Table 2, and Figure 3A shows the predicted probabilities of the model as well as the ROC analysis.

**Table 2:**
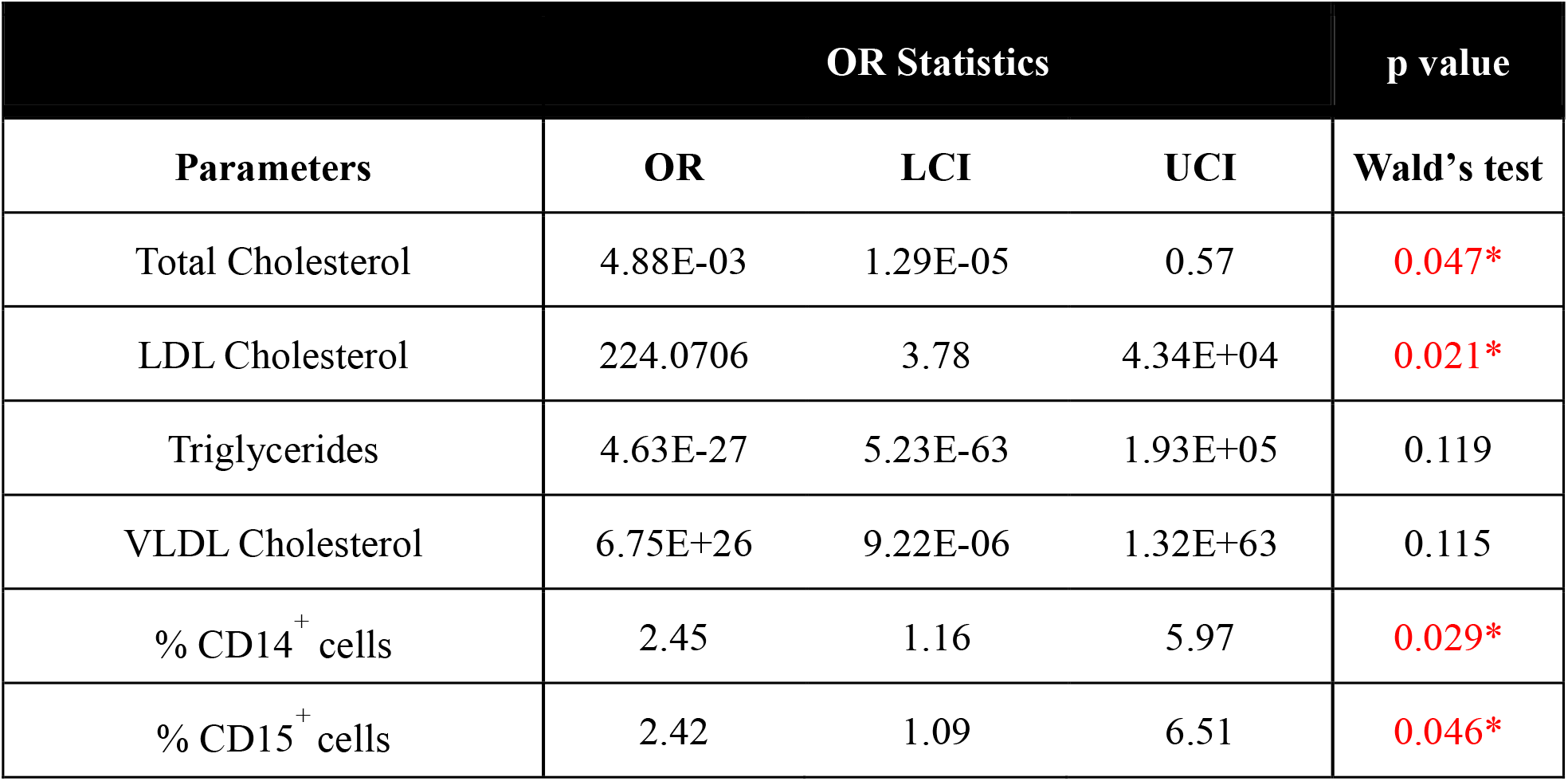
Summary of multivariable Biochemical logistic regression model. OR indicates Odds ratio and LCI and UCI indicate lower and upper confidence interval, respectively. Wald’s test was used to assess significance of predictors contributing to model’s performance. * indicates significance with p < 0.05

**Figure 3:**
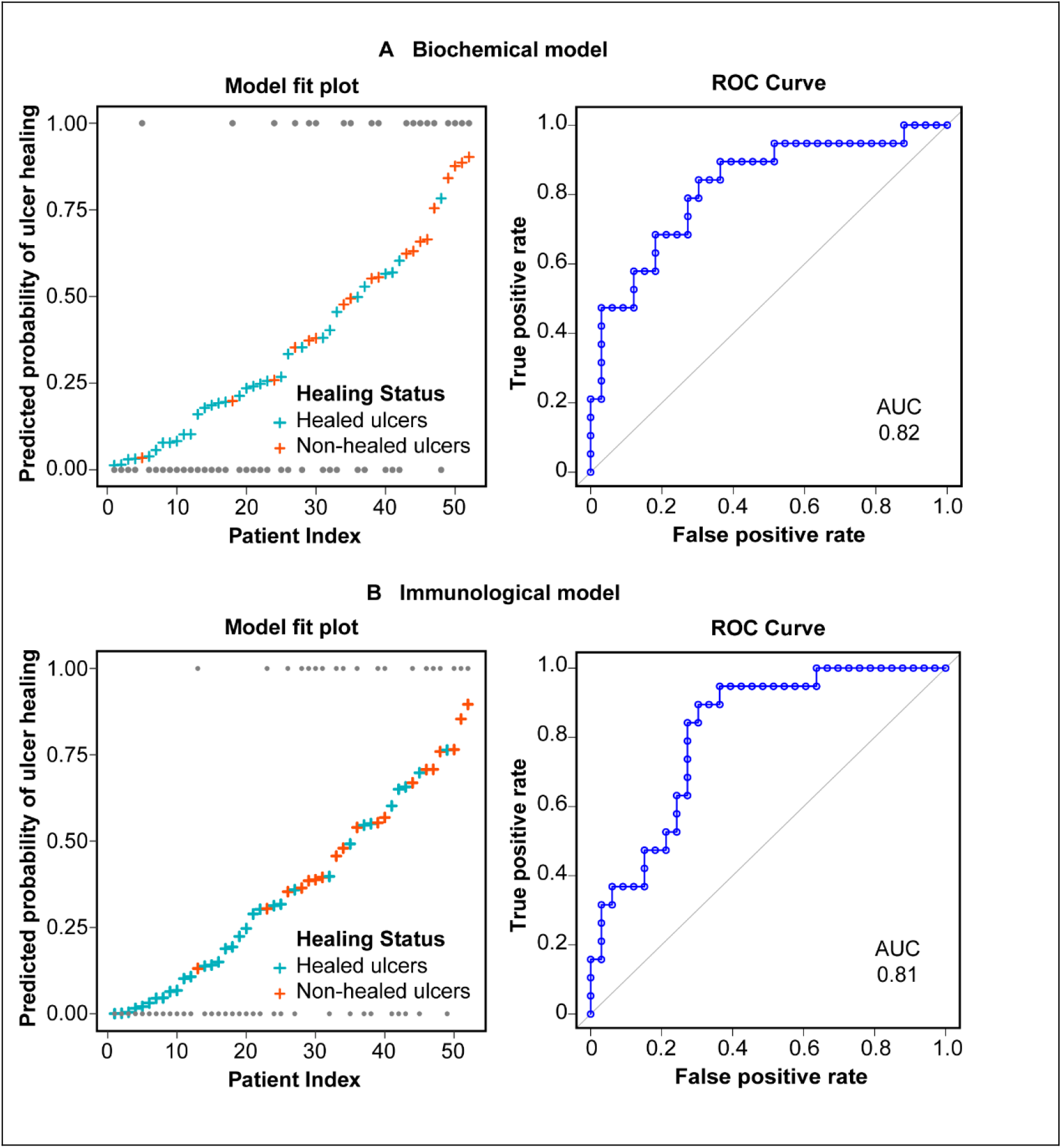
Multivariable Logistic Regression. A and B –. Biochemical and cellular parameters **(A)** and Immunological parameters **(B)** were used to predict probabilities of healing. The graphs on the left show the fit of the model with reference probabilities indicated as grey dots. The graphs on the right show the receiver operating characteristics (ROC) analysis with area under the curve (AUC) measurements.

Similarly, a stepwise feature selection identified four of 15 immunological phenotypic parameters to build the Immunological model, three of which were found to be significant according to Wald’s test (Table 3). Chi-square and HL GOF tests showed p value 0.002 and 0.368, respectively indicating that the model is statistically significant, and the AIC was found to be 61.21. Odd’s ratio and corresponding CI are summarized in Table 3. Figure 3B shows the predicted probabilities of the model as well as the ROC analysis.

**Table 3:**
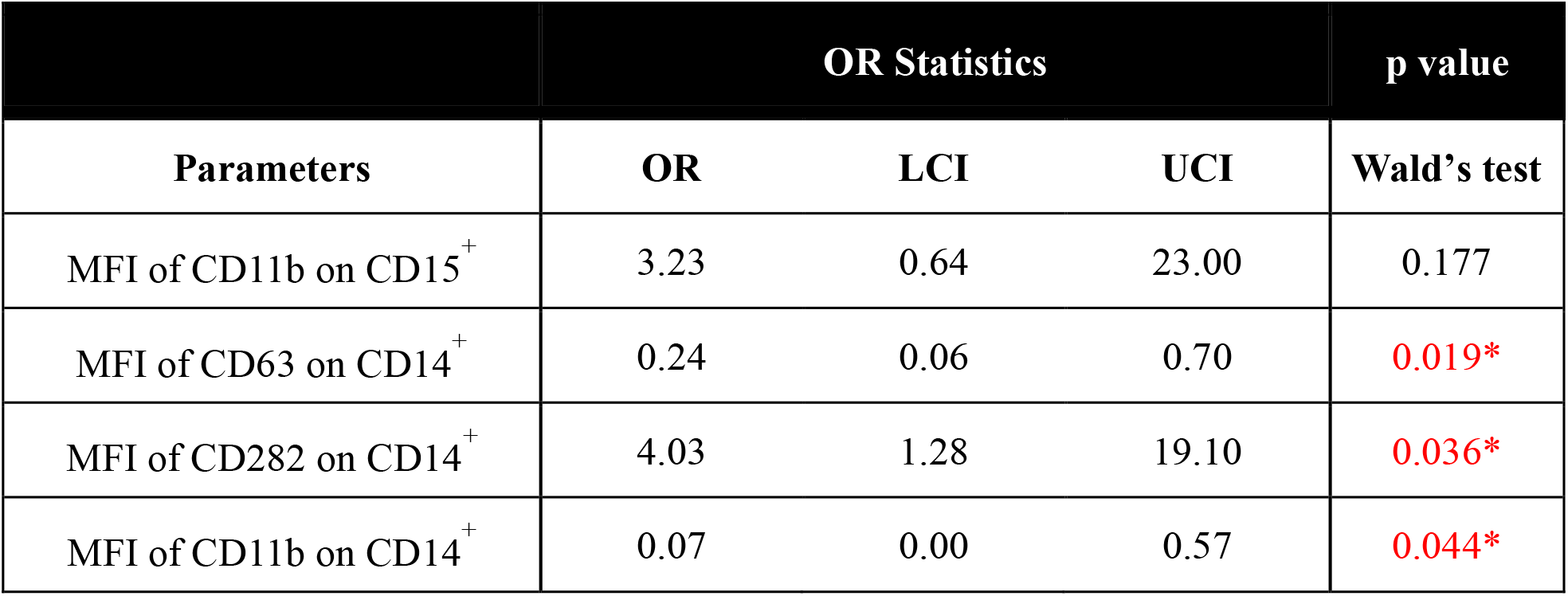
Summary of multivariable Immunological logistic regression model. OR indicates Odds ratio and LCI and UCI indicate lower and upper confidence interval, respectively. Wald’s test was used to assess significance of predictors contributing to model’s performance. * indicates significance with p < 0.05

| Parameters | OR Statistics |  |  | p value |
| --- | --- | --- | --- | --- |
|  | OR | LCI | UCI | Wald's test |
| MFI of CD11b on CD15<sup>+</sup> | 3.23 | 0.64 | 23.00 | 0.177 |
| MFI of CD63 on CD14<sup>+</sup> | 0.24 | 0.06 | 0.70 | 0.019* |
| MFI of CD282 on CD14<sup>+</sup> | 4.03 | 1.28 | 19.10 | 0.036* |
| MFI of CD11b on CD14<sup>+</sup> | 0.07 | 0.00 | 0.57 | 0.044* |

While both the biochemical and immunological models of multivariable logistic regression showed that specific groups of parameters could help to predict non-healing, we were interested in determining if combining the parameters would enhance model performance. For such an analysis, significant parameters of both the multivariable models were chosen and a stepwise feature selection was performed to retain only salient parameters in the model. This new model, with four parameters, showed an excellent fit (Figure 4A), an improved AUC of 0.87 in the ROC analysis (Figure 4B), and a lowered AIC score of 56.25. The Chi-square and HL GOF test show p value 0.0002 and 0.3086, respectively indicating that the model is statistically significant. Two (LDL cholesterol and CD63 MFI among monocytes) of the four selected parameters showed significance as assessed by Wald’s test (Table 4). LDL cholesterol showed an OR 18.83 (CI 18.83-342) indicating positive association with healing outcome, while the MFI of CD63 among monocytes (CD14^+^) showed an OR 0.12 (CI 0.02-0.45) suggesting a negative association with healing outcome. Together, these metrics suggest that the combined model performed the best in predicting non-healing among individuals with early-stage diabetic foot ulcers.

**Table 4:** Summary of multivariable combined logistic regression model. OR indicates Odds ratio and LCI and UCI indicate lower and upper confidence interval, respectively. Wald’s test was used to assess significance of predictors contributing to model’s performance. * indicates significance with p < 0.05 ** indicate p < 0.01

| Parameters | OR Statistics |  |  | p value |
| --- | --- | --- | --- | --- |
|  | OR | LCI | UCI | Wald's test |
| Total Cholesterol | 0.15 | 0.01 | 1.07 | 0.105 |
| LDL Cholesterol | 18.83 | 2.22 | 342.00 | 0.021* |
| % CD14<sup>+</sup> cells | 2.18 | 1.03 | 5.40 | 0.057 |
| MFI of CD63 on CD14<sup>+</sup> | 0.12 | 0.02 | 0.45 | 0.006** |

**Figure 4:**
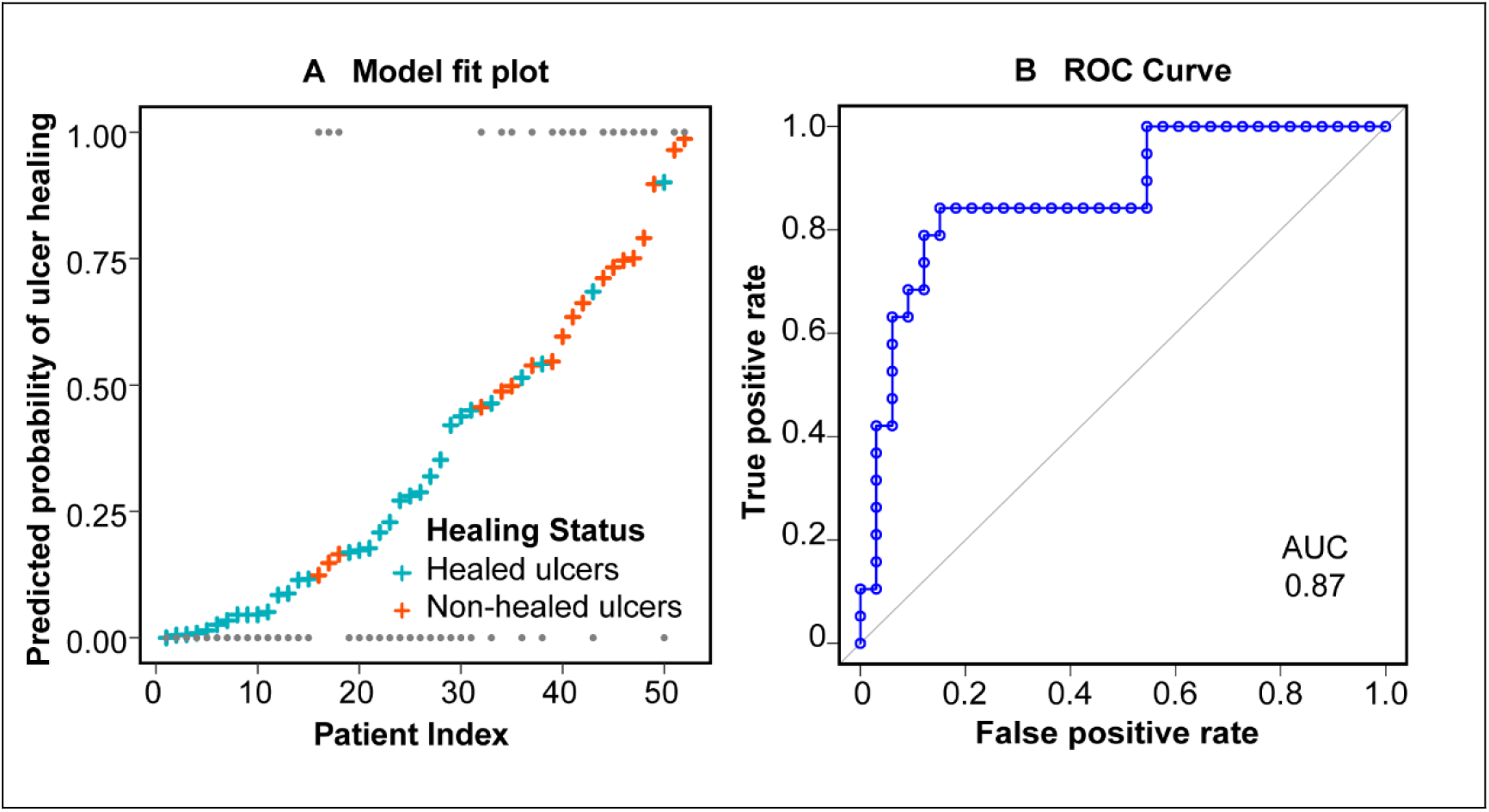
Multivariable Combined logistic regression model performance. **A** – A representation of the predicted probabilities of a model, which combines both biochemical and immunological parameters, against reference probabilities (shown by grey dots). **B** – Receiver operating characteristics (ROC) analysis of combined model shows that the area under the curve (AUC).

## DISCUSSION

The goal of our study was to identify clinically measurable parameters that could help predict individuals in whom early-stage DFUs fail to heal. Hence, we focused on collecting and analyzing histopathological, biochemical, and immunological parameters that are relatively easy to collect in many health-centers. Our analysis revealed that the histopathological data did not correlate with healing, and no individual biochemical or immunological parameter was a good predictor of non-healing. However, multivariable logistic regression analysis using a combination of biochemical and immunological parameters was able to predict non-healing. Specifically, the combined analysis determined that serum LDL cholesterol (OR 18.83, CI 2.22 and 342.00) and the expression level of the protein CD63 among monocytes (OR 0.12, CI 0.02 and 0.45) were significant predictors of non-healing in early-stage DFUs.

Previous cohort studies on individuals with DFU have identified systemic inflammatory mediators that appear to correlate with inflammation at the wound site and non-healing. For example, Veves and colleagues report the presence of increased serum levels of TNF-α, FGF-2, MCP-1 and MMP-9 in individuals who developed DFU eight months after the serum measurements and failed to heal by 12 weeks(18). Recently, the same authors identified unique transcriptomic signatures in individuals with foot ulcers and those that do not heal(26). Other studies also show that neutrophil elastase and citrullinated H3, markers of neutrophils extracellular traps (NETs), were observed to be increased in plasma of non-healed DFU individuals compared to healed individuals(14,27). In terms of diabetic wound-specific biomarkers for healing, independent studies by Wong et al, Fadini et al and Yang et al observed an increased presence of NET components in wounds of diabetic mice and in individuals with DFU thereby correlating NETosis to delayed healing (13,14,27). While these studies clearly establish that both local and systemic inflammation correlate with poor healing, measurement of the identified mediators are often difficult due to the labor-intensive methodologies involved or the requirement for equipment that are not yet available in many clinical centers.

Facile measurable parameters such as wound characteristics (wound size and area) have also been reported as predictors of non-healing. Sheehan et al. showed that the percentage change in wound area after four weeks was a robust predictor of non-healing observed at twelve weeks(7). Further, Lavery et al. demonstrated that percentage reduction in wound area by one week was a strong predictor for non-healing at sixteen weeks(8). However, these observations were from patients with late stage or amputated wounds, which may not be applicable to early-stage wounds. Additionally, precise cutoffs of wound dimension changes are also difficult to establish, as they may vary between study cohorts, ulcer type, and stage of wounds. In this context, serum biochemical indicators that are routinely measured in the clinic and immunological phenotyping (which is becoming common in many tertiary care centers) are easy-to-use as potential predictors of wound healing.

The major strength of our study is the identification of such measurable biochemical and immunological parameters as predictors of non-healing in early-stage DFU individuals. Our study shows that LDL cholesterol among biochemical parameters and expression level of protein CD63 on monocytes among immunological parameters (along with total cholesterol and percentage of monocytes), together, serve as strong predictors of non-healing in individuals with active foot ulcers. LDL cholesterol has been implicated in inflammation and dysregulated innate immune cell function(28). Hence, individuals with increased levels of LDL cholesterol may have systemic low-grade inflammation, which might impact many tissues, including DFUs. Additionally, LDL cholesterol has been shown to have inhibitory effect on endothelial cell proliferation, and has been shown to cause delays in wound healing in mouse models of research(29). In concurrence with these observations, our analysis reveals that higher levels of LDL cholesterol correlate with non-healing of early-stage foot ulcers, possibly through the establishment of higher systemic-inflammatory levels and lowered endothelial cell proliferation.

The protein CD63 is known to be an important component of phagosomes(30) and plays a role in trafficking of proteins from the membrane into the cell (31). Additionally, upon maturation of monocytes, surface expression of CD63 is known to decrease(32). Monocytes in circulation are known to be recruited to the wound bed where they differentiate into macrophages releasing pro and anti-inflammatory mediators to accelerate healing(33). While the exact link between CD63 and monocyte activity in wound healing (or microbial killing) is not clear, we speculate that a circulating monocyte with lower surface CD63 expression might indicate matured cell with altered phenotype or a cell with reduced ability to engage in wound healing. Such a state could explain the inverse relationship between monocytic CD63 expression and healing of diabetic wounds.

In diabetic individuals, hyperglycemia is known to play a role in establishing and maintaining inflammation (34–36). In that context, a surprising observation in our study is the lack of correlation between hyperglycemia (both HbA1c and fasting blood sugar) and non-healing. This observation is in contrast to a decade old retrospective study by Christman et al., which showed that higher HbA1c strongly correlates to poor healing in DFU individuals(37). Two possible reasons for the difference between our observations and this study, is the stage of ulcer under consideration (we focus on early-stage ulcers compared to aggressive late stage or amputated wounds in the other study) and the method used to classify healing (we measure healing at an endpoint compared to rate of healing used in the other study). However, our data and observations are in line with a meta-analysis performed on individuals with neuropathic diabetic wounds(38) and a more recent retrospective study on individuals with DFU (39). Based on these studies and our observations, it does appear that while hyperglycemia may play a role in the development of foot ulcers, it does not correlate with healing status.

One of the major limitations of this study is the relatively small sample size of the cohort, which may limit the statistical power. Nevertheless, our observations provide new information on measurable predictors for non-healing. Second is the classification system used for assessing healing status, which was set as 50% reduction in wound area by 4 weeks following thresholds reported by Sheehan et al. and Lavery et al. (7,8). Due to challenges associated with following-up patients in our setting, we were unable to physically verify if the individuals classified as healed showed complete wound healing by ∼16 weeks. Third, the duration that the individual has had an ulcer for before presentation in the clinic was determined through an oral conversation but could not be verified as in many cases no prior medical records exist. Lastly, many of the individuals who visit this center have uncontrolled diabetes, and while this may be common for many government health centers in India, it may not be the norm elsewhere.

## CONCLUSION

In summary, our cross-sectional cohort study analyzing clinical, biochemical, and immunological parameters among individuals with early-stage DFU showed that no single parameter correlates effectively with non-healing of wounds. However, a multivariable logistic regression model of the data suggests that LDL cholesterol and the expression level of CD63 on monocytes are strong and significant predictors of non-healing, when used along with total cholesterol and percentage of monocytes. While validation studies in larger cohorts are required, our observations have the potential to aid clinicians in identifying individuals at the risk of a poor healing outcome, and hence pursue more aggressive or alternate treatment strategies.

## Data Availability

All data produced are available online at https://github.com/Immunoengineeringlab/DFU_IISc_KIER_JVR_Data.

## ACKNOWLEDGMENTS

The authors acknowledge support from the Rajiv Gandhi University of Health Sciences, Govt. of Karnataka (17C008B). This study was also funded in part by the Biodesign and Bioengineering Initiative (Phase II), Department of Biotechnology, Govt. of India. It was also funded by the Dr. Vijaya and Rajagopal Rao laboratory for Biomedical Engineering at IISc. The authors have no conflict of interest to declare. JVR helped design the study, collected the immunological data, analyzed all data, and wrote the manuscript. SK and PA assisted with immunological data collection. VKD and RDS helped with subject recruitment and assisted with biopsy as well as immunological sample processing. CHC and PB assisted with study design, recruited subjects, collected the biopsies, and assisted with writing the manuscript. KSR helped with study design, performed the histopathological analysis and assisted with writing the manuscript. SJ designed the study, assisted with data collection and analysis, and wrote the manuscript.

