## Supplementary for "Biochemical and Immunological Predictors of Non-healing in Individuals with Early-stage Diabetic Foot Ulcers"

### Table of Contents

Supplementary Figures 2 – 6

Supplementary Tables 7 – 10

### Supplementary Figures

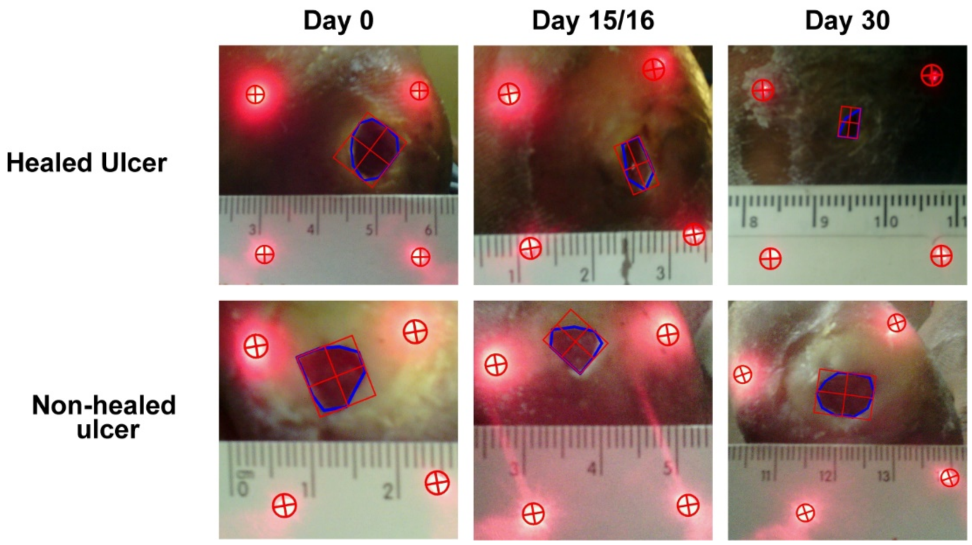


**Supplementary Figure 1: Representative images of diabetic foot ulcers.** Images were captured using a near infra-red camera (WoundZoom) over a one-month follow-up window. These images were used to calculate percentage reduction in wound area using ImageJ. Individuals were classified as healed if the wound area reduced to >50% (of the area on day of recruitment) on Day 30.

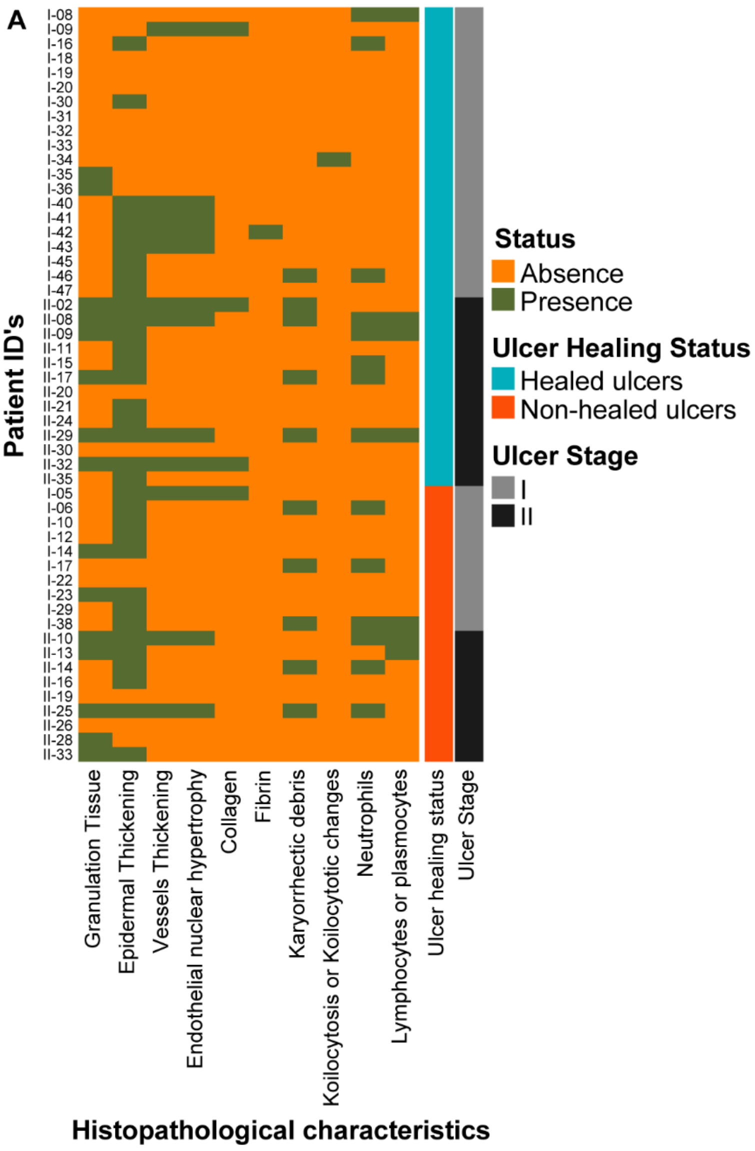


**B**

| | Healing status | Ulcer stage |
|---|---|---|
| **Parameters** | **p value** | **p value** |
| **Granulation tissue** | 0.36 | 0.01* |
| **Epidermal Thickening** | 0.38 | 0.15 |
| **Vessel Thickening** | 0.50 | 0.74 |
| **Endothelial nuclear hypertrophy** | 0.50 | 0.74 |
| **Collagen** | 1.00 | 1.00 |
| **Fibrin** | 1.00 | 1.00 |
| **Karyorrhectic debri** | 0.47 | 0.29 |
| **Koilocytosis** | 1.00 | 1.00 |
| **Ulcer stage** | 0.77 | - |
| **Neutrophils** | 0.75 | 0.22 |
| **Lymphocytes** | 0.70 | 0.12 |

**Supplementary Figure 2: Histopathological analyses of wound biopsy samples. A -** Heatmap of histopathological data. Green indicates presence and orange indicates absence of measured parameter in each individual. **B** - Fisher's exact test for assessment of correlation between histopathological variables and healing outcome or ulcer stage. Presence of granulation tissue was the only parameter found to correlate to stage of ulcer, and none correlated with healing status. * indicates significance with $p < 0.05$

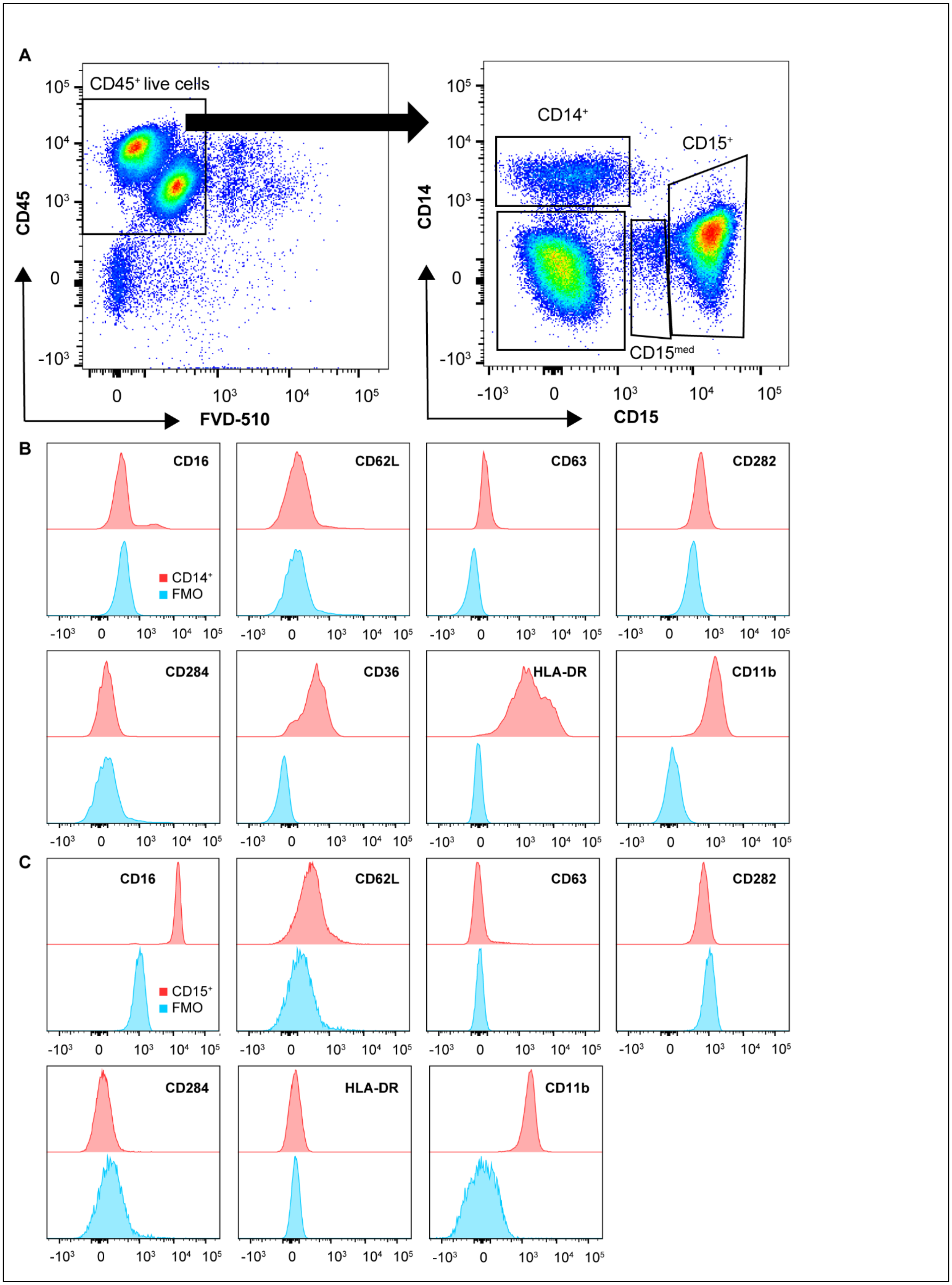
A
CD45⁺ live cells
CD45
FVD-510
CD14⁺
CD15⁺
CD15med
CD14
CD15
B
CD16
CD62L
CD63
CD282
CD284
CD36
HLA-DR
CD11b
CD14⁺
FMO
C
CD16
CD62L
CD63
CD282
CD284
HLA-DR
CD11b
CD15⁺
FMO

**Supplementary Figure 3: Gating strategy to identify neutrophils and monocytes through flow cytometry. A** – Among $CD45^+$ live cells neutrophils were identified as $CD15^+$ and monocytes as $CD14^+$ ($CD15^-$) cells. **B** and **C** - Representative histograms showing expression levels of all assessed markers expressed on monocytes (B) and neutrophils (C) along with corresponding FMOs (blue)

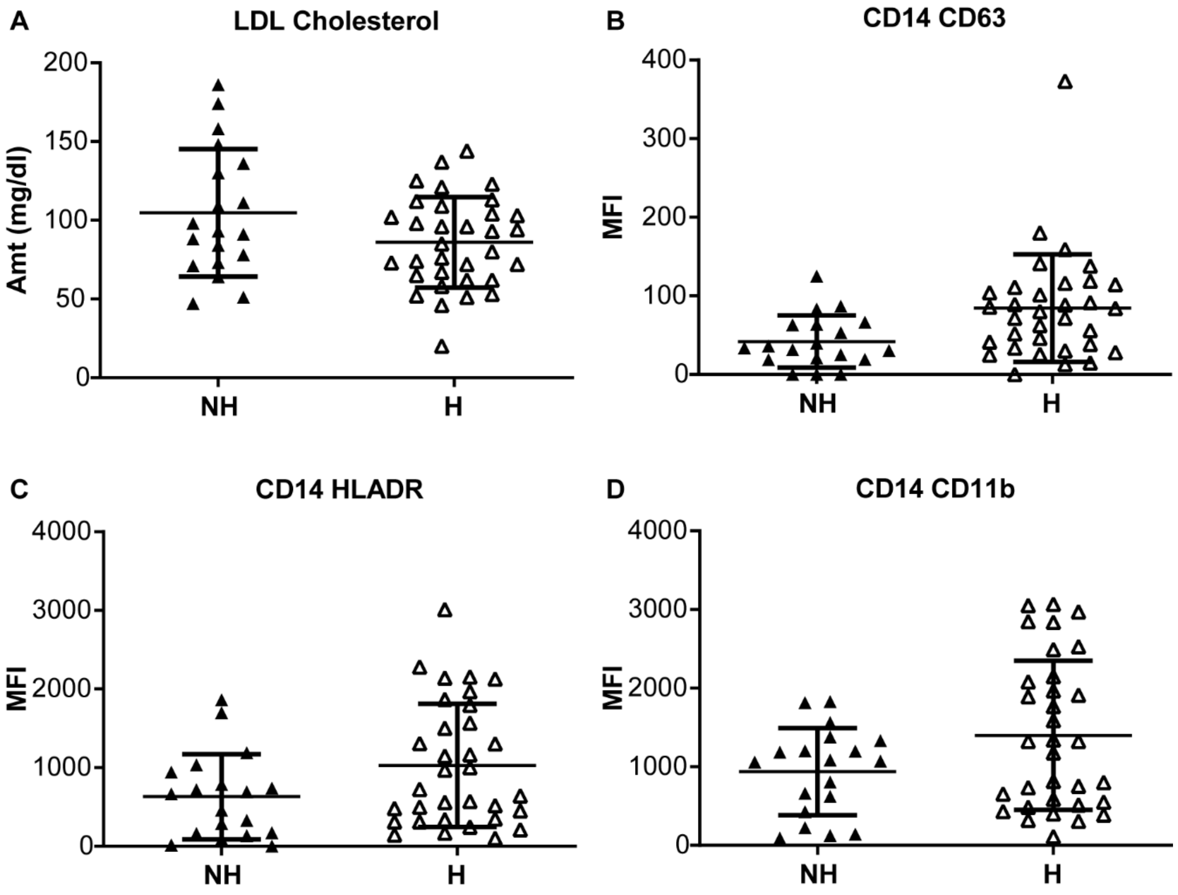


**Supplementary Figure 4: Univariate analysis of biochemical and immunological parameters.** Values of **A** - LDL cholesterol, **B** - MFI of CD63, **C** - MFI of HLA-DR, and **D** - MFI of CD11b (immune markers on monocytes) among non-healed (NH) and healed (H) individuals are presented. Student's t test with Welch's correction was used to determine statistical significance. * indicates significance with $p < 0.05$

### Supplementary Tables

| Fluorophore | Panel 1 | Panel 2 |
|---|---|---|
| Brilliant Violet 510 | Live-Dead | |
| Brilliant Violet 605 | CD14 (M5E2) | |
| Brilliant Violet 650 | CD15 (HI98) | |
| Brilliant Violet 786 | CD62L (SK11) | CD284 (TF901) |
| FITC / Alexa Fluor 488 | CD16 (3G8) | CD282 (11G7) |
| PE | CD63 (H5C6) | CD36 (CB38) |
| PE-CF594 | - | HLA-DR (G46-6) |
| APC / Alexa Fluor 647 | CD66b (G10F5) | - |
| Alexa Fluor 700 | CD11b (ICRF44) | |
| APC-Cy7 | CD45 (2D1) | |

**Supplementary Table 1: Antibody panel used for immunophenotyping of neutrophils and monocytes isolated from blood.** The specific clone is indicated in parenthesis

| Parameters | p value |
|---|---|
| Age | 0.820 |
| BMI | 0.301 |
| HbA1c | 0.959 |
| FBS | 0.256 |
| PPBS | 0.670 |
| Total Cholesterol | 0.314 |
| HDL Cholesterol | 0.948 |
| LDL Cholesterol | 0.087 |
| S Triglycerides | 0.394 |
| VLDL Cholesterol | 0.398 |
| Total Chol/HDL Chol | 0.239 |
| ESR | 0.660 |
| Alkaline Phosphatase | 0.879 |
| % $CD14^+$ cells | 0.246 |
| % $CD15^+$ cells | 0.287 |
| % $CD15^{med}$ cells | 0.157 |

| Parameters | p value |
|---|---|
| MFI of CD16 on $CD15^+$ | 0.752 |
| MFI of CD62L on $CD15^+$ | 0.161 |
| MFI of CD63 on $CD15^+$ | 0.059 |
| MFI of CD282 on $CD15^+$ | 0.687 |
| MFI of CD284 on $CD15^+$ | 0.922 |
| MFI of HLA-DR on $CD15^+$ | 0.427 |
| MFI of CD11b on $CD15^+$ | 0.295 |
| MFI of CD16 on $CD14^+$ | 0.571 |
| MFI of CD62L on $CD14^+$ | 0.382 |
| MFI of CD63 on $CD14^+$ | 0.004** |
| MFI of CD282 on $CD14^+$ | 0.770 |
| MFI of CD284 on $CD14^+$ | 0.989 |
| MFI of CD36 on $CD14^+$ | 0.123 |
| MFI of HLA-DR on $CD14^+$ | 0.035* |
| MFI of CD11b on $CD14^+$ | 0.031* |

**Supplementary Table 2: Univariate analysis of clinical, biochemical and immunological parameters.** P values obtained from statistical analysis (Student's t-test with Welch's correction) that compares healed to non-healed individuals is presented in the table. * indicates significance with $p < 0.05$, ** indicate $p < 0.01$

| Parameters | AUC |
|---|---|
| Age | 0.51 |
| BMI | 0.61 |
| HbA1c | 0.50 |
| FBS | 0.60 |
| PPBS | 0.53 |
| Total Cholesterol | 0.57 |
| HDL Cholesterol | 0.53 |
| LDL Cholesterol | 0.61 |
| Triglycerides | 0.56 |
| VLDL Cholesterol | 0.55 |
| Total Chol/HDL Chol | 0.59 |
| ESR | 0.50 |
| Alkaline Phosphatase | 0.52 |
| % $CD14^+$ cells | 0.60 |
| % $CD15^+$ cells | 0.59 |
| % $CD15^{med}$ cells | 0.52 |

| Parameters | AUC |
|---|---|
| MFI of CD16 on $CD15^+$ | 0.51 |
| MFI of CD62L on $CD15^+$ | 0.62 |
| MFI of CD63 on $CD15^+$ | 0.53 |
| MFI of CD282 on $CD15^+$ | 0.60 |
| MFI of CD284 on $CD15^+$ | 0.61 |
| MFI of HLA-DR on $CD15^+$ | 0.53 |
| MFI of CD11b on $CD15^+$ | 0.54 |
| MFI of CD16 on $CD14^+$ | 0.60 |
| MFI of CD62L on $CD14^+$ | 0.56 |
| MFI of CD63 on $CD14^+$ | **0.73** |
| MFI of CD282 on $CD14^+$ | 0.50 |
| MFI of CD284 on $CD14^+$ | 0.58 |
| MFI of CD36 on $CD14^+$ | 0.63 |
| MFI of HLA-DR on $CD14^+$ | 0.65 |
| MFI of CD11b on $CD14^+$ | 0.63 |

**Supplementary Table 3: Summary of Receiver operating characteristics of all biochemical and immunological parameters.** Area under the curve (AUC) is tabulated for all parameters

| | OR Stats | | | p value | | ROC Analysis |
|---|---|---|---|---|---|---|
| **Parameters** | **OR** | **LCI** | **UCI** | **Wald's test** | **Chi square test** | **AUC** |
| MFI of $CD63$ on $CD14^{+}$ | 0.26 | 0.08 | 0.68 | 0.013* | 0.003** | 0.73 |
| MFI of HLA-DR on $CD14^{+}$ | 0.52 | 0.24 | 0.98 | 0.064 | 0.044* | 0.65 |
| MFI of CD11b on $CD14^{+}$ | 0.53 | 0.26 | 1.00 | 0.066 | 0.049 | 0.63 |

**Supplementary Table 4: Summary of univariate logistic regression model.** OR indicates odd's ratio calculated for each parameter. LCI and UCI indicate lower and upper confidence interval, respectively. AUC indicates area under the ROC curve. * indicates significance with $p < 0.05$, ** indicate $p < 0.01$
